# Recovery of clinical, cognitive and cortical activity measures following mild traumatic brain injury (mTBI): a longitudinal investigation

**DOI:** 10.1101/2022.06.03.22275984

**Authors:** Hannah L. Coyle, Neil W. Bailey, Jennie Ponsford, Kate E. Hoy

**Affiliations:** Central Clinical School Department of Psychiatry, Monash University, Melbourne, Victoria, Australia; Epworth Centre for Innovation in Mental Health, Epworth Healthcare and Department of Psychiatry, Monash University, Camberwell, Victoria, Australia; Turner Institute for Brain and Mental Health, Monash University; Monash-Epworth Rehabilitation Research Centre, Epworth Healthcare, Melbourne, Australia

**Author notes:** Corresponding author: Prof Kate Hoy, Central Clinical School Department of Psychiatry, Monash University, Melbourne, Australia. These authors contributed equally (Hannah L. Coyle, Neil W. Bailey).

**Keywords:** cognition, electroencephalography (EEG), mild traumatic brain injury (mTBI), post-concussion symptoms, transcranial magnetic stimulation (TMS)

## Abstract

The mechanisms that underpin recovery following mild traumatic brain injury (mTBI) remain poorly understood. Identifying neurophysiological markers and their functional significance is necessary to develop diagnostic and prognostic indicators of recovery. The current study assessed 30 participants in the subacute phase of mTBI (10-31 days post-injury) and 28 demographically matched controls. Participants also completed 3 month (mTBI: N = 21, control: N = 25) and 6 month (mTBI: N = 15, control: N = 25) follow up sessions to track recovery. At each time point, a battery of clinical, cognitive, and neurophysiological assessments was completed. Neurophysiological measures included resting-state electroencephalography (EEG) and transcranial magnetic stimulation combined with EEG (TMS-EEG). Outcome measures were analysed using mixed linear models (MLM). Group differences in mood, post-concussion symptoms and resting-state EEG resolved by 3 months, and recovery was maintained at 6 months. On TMS-EEG derived neurophysiological measures of cortical reactivity, group differences ameliorated at 3 months but re-emerged at 6 months, while on measures of fatigue, group differences persisted across all time points. Persistent neurophysiological changes and greater fatigue in the absence of measurable cognitive impairment may suggest the impact of mTBI on neuronal communication may leads to increased neural effort to maintain efficient function. Neurophysiological measures to track recovery may help identify both temporally optimal windows and therapeutic targets for the development of new treatments in mTBI.

## 1. Introduction

Mild traumatic brain injuries (mTBI’s) are associated with a wide range of cognitive, behavioural and affective symptoms (Mass et al., 2017). mTBI pathophysiology is associated with symptoms via diffuse and dynamic processes (Giza et al., 2014). White matter tracts (bundles of axons that connect neurons in distal brain regions into functional circuits) are particularly vulnerable to a complex spectrum of axonal changes, known as diffuse axonal injury (DAI) (Armstrong et al., 2016). Another important cellular mechanism affected is the excitation-inhibition (E:I) balance, the ratio of excitatory to inhibitory inputs in a neural circuit which contributes to maintaining neural firing and induction of plasticity (Eichler et al., 2008). Levels of glutamate (the primary excitatory) and GABA (the primary inhibitory) neurotransmitter are linked to homeostatic control of the E:I balance. Crucially, this balance is considered to underpin induction of plasticity (Froemke et al., 2015) and its dysregulation has been linked to cognitive, behavioural and affective symptoms across neurological and neuropsychiatric disorders (Kegeles et al., 2012; Rubenstein et al., 2003; Yizhar et al., 2011).

Historically, symptoms were thought to be transient after mTBI, with return to pre-injury functioning occurring within 90 days (Rohling et al., 2011). More recent research has reported the presence of persistent symptoms in a subgroup of individuals months to years’ post-injury (McInnes et al., 2017). Variability is likely influenced by the dynamic interaction of the initial injury with an individual’s neurobiology at the time of injury and across their recovery trajectory, making the evolution of each mTBI potentially unique (Coyle et al., 2018). Developments in neuroimaging have sought to characterise these trajectories, and structural and functional connectivity changes have been demonstrated in acute and chronic mTBI (Eierud et al., 2014; Medeglia et al., 2017). Associations between symptoms and network connectivity changes have also been demonstrated; however heterogeneity remains a significant hurdle to the development of diagnostic and prognostic indicators of recovery (Coyle et al., 2018). E:I balance dysregulation has also been proposed to contribute to persistent symptoms in mTBI (Guerriero, et al., 2015). Linking these neurophysiological changes with functional consequences over time will help to clarify heterogeneous recovery trajectories, which will potentially assist with identifying individuals at risk of persistent symptoms.

Electroencephalography (EEG) measures of cortical activity have high temporal resolution and are well placed to probe pathophysiology following mTBI. Recording EEG at rest and during cognitive tasks provides information on cortical activity during passive and active information processing and can detect subclinical abnormalities in brain activity related to cognition (Rapp et al., 2015). Impaired coherence (a measure of functional connectivity) has also been demonstrated following mTBI during verbal and visuo-spatial Sternberg tasks during working memory (WM), but not at rest (Kumar et al., 2009). However, cross-sectional designs and significant variability in mean time since injury (ranging between 45 days and 10 years’ post-injury) limit the temporal specificity and generalisability of findings.

TMS combined with EEG is another powerful way to examine brain function. TMS is a form of non-invasive brain stimulation that can be used to measure cortical activity. In the case of TMS-EEG, the application of TMS to the scalp generates a brief perturbation in neural activity which is detected in the EEG activity – this is referred to as a TMS Evoked Potential (TEP). The spread of TEP activation across the scalp provides information about cortical reactivity. Characteristics of TEP’s such as amplitude, latency and polarity suggest time locked properties of underlying mechanisms. For example, the P60 and N100 components have been associated with activation of GABAergic mechanisms (Premoli et al. 2014; Rogasch et al. 2013, Rogasch et al. 2015, Belardinelli et al. 2021, Kaarre et al. 2018, Noda 2020), as well as somatosensory and auditory processing of the TMS pulse (Nikouline et al. 1999, Conde et al. 2019, Biabani et al. 2019, Biabani et al. 2021). To date, only TMS measures from the motor cortex, referred to as motor evoked potentials (MEPs), have been assessed using a longitudinal design in mTBI. Greater intra-cortical inhibition, considered to be indicative of a GABAergic response, was reported at 72-hours and 2 months’ post mTBI (Edwards et al., 2017; Miller et al., 2014). Although MEPs provide valuable information on corticospinal excitability and motor function, the diffuse and dynamic nature of mTBI (with predominately a clinical and cognitive symptom profile) highlights the importance of exploring cortical activity outside of the motor cortex in clinically and cognitively relevant brain regions. To summarise, the major limitations of previous EEG and TMS-EEG research in mTBI include; primarily cross-sectional designs, significant variations in time since injury, and application of TMS solely to the motor cortex.

A comprehensive characterisation of post-injury symptoms, function, neurophysiology, and repeated measurements across time are necessary to improve our understanding of mechanisms of recovery and interactions between pathophysiology and symptoms following mTBI. To address this, we designed a prospective, longitudinal, controlled cohort study that assessed clinical, cognitive and neural measures in individuals <1 month post mTBI (referred to as the sub-acute time point) and at 3 month and 6 month follow up time points. A demographically matched group of control participants with no history of head injury completed the same protocol. Between-group comparisons at the sub-acute timepoint (< 4 weeks) have been reported previously (Coyle et al., 2022). In brief, mTBI participants reported higher rates of mood, fatigue and post-concussive symptoms than controls, as well as reduced performance on a test of verbal learning. Neurophysiological findings included mTBI participants having greater alpha power at rest and the TMS-EEG findings showed that mTBI participants demonstrated smaller P60 amplitudes and greater N100 amplitudes compared to controls.

In the present study, we report results from 3 and 6 month follow up time points for the measures that were significantly different between mTBI and controls at 4 weeks’ post mTBI. The aim was to comprehensively explore recovery by tracking clinical, cognitive and neurophysiological changes at 3- and 6-months’ post-injury (referred to in this study as T1 and T2 respectively). Based on past research demonstrating a relationship between symptoms and pathophysiology, we hypothesised that persistent symptoms will be accompanied by persistent pathophysiological changes. To our knowledge this is the first investigation of TMS-EEG in a longitudinal design in mTBI.

## 2. Material and Methods

A total of 58 participants were recruited (30 mTBI, 28 controls). The 30 participants with mTBI were recruited from the Emergency Department and trauma wards of the Alfred Hospital, Melbourne (mean days since injury = 19.70, SD = 16.96, range 10-31, mean age at injury = 35.43 years, *SD* = 10.31). 28 demographically matched controls with no history of TBI (mean age = 31.65 years, *SD* = 9.06) were also recruited. No participants had a history of seizures, psychiatric or neurological illnesses, unstable medical conditions, were pregnant or taking prescribed medication known to directly or significantly influence EEG findings. mTBI was classified as exhibiting an initial Glasgow Coma Scale (GCS) score of 13-15, loss of consciousness < 30 minutes and post-traumatic amnesia (PTA) < 24 hours (Carroll et al., 2004). mTBI injury characteristics are presented in Supplementary Table 1, and demographic and clinical characteristics are shown in Supplementary Table 2. All participants provided written informed consent prior to commencement of study procedures. The study received approval from both Alfred Health and Monash University Ethics Committees.

### 2.1 Procedure

To investigate the time course of recovery following mTBI, clinical, cognitive and neural assessments were completed in the sub-acute phase, (i.e. within 4 weeks post-injury), referred to as the sub-acute time point, and at 3 month follow up and a 6 month follow up time points. Control participants completed the same sequence of assessments to enable comparison between the two groups across all time points. See Supplementary Table 1 and Supplementary Figure 1 for more information on participants who completed each timepoint. In order to focus the analyses conducted from this substantial data set, our apriori data analysis plan was to only include measures that were identified to be dysregulated (i.e. significantly differentiated the groups at the initial sub-acute assessment). Measures are described in detail below.

### 2.2 Measures

#### 2.2.1 Clinical and Cognitive Measures

Clinical measures assessed mood (Hospital Anxiety and Depression scale: HADS) (Zigmond et al., 1983), fatigue (Multidimensional Fatigue Inventory: MFI) (Smets et al., 1995) and post concussive symptoms (Rivermead Post Concussive Symptom Questionnaire: RPQ, Rivermead Head Injury Follow-Up Questionnaire: RHFUQ) (Crawford et al., 1996; King et al., 1995). The sub-acute study results showed that mTBI participants reported greater symptom severity on all clinical measures and so all were examined here (Coyle et al., 2022). In the current analyses data from domains of the MFI (General Fatigue, Physical Fatigue, Mental Fatigue, Reduced Motivation and Reduced Activity) and HADS (Anxiety symptoms and Depressive symptoms) were individually summed to create total fatigue and total mood scores at each time point. This was completed to reduce the number of outcome variables and multiple comparisons. As the RPQ (16-item) and RHFUQ (10-item) are on different scales, a total percentage severity score was calculated to standardise these measures. Cognitive measures included; Wechsler Test of Adult Reading (WTAR) as a measure of pre-morbid IQ, Trail Making Test A and B (TMT-A, TMT-B), subtests from the Wechsler Adult Intelligence Scale-IV (WAIS-IV) Working Memory Index (WMI) and Processing Speed Index (PSI), Brief Visual Memory Test (BVMT), Rey Auditory Verbal Learning Test (RAVLT) and Controlled Word Association Test (COWAT). For memory measures, the RAVLT and BVMT, alternative forms were used at each time point. At the sub-acute time point, after controlling for pre-morbid IQ, group differences were only demonstrated on total words recalled on trial 1 (T1) of the RAVLT (Coyle et al., 2022), so this was the only cognitive measure included in the current analyses.

#### 2.2.2 Neural Measures

Neural measures included resting EEG and TMS-EEG. Task related EEG was also collected during a working memory task, however there were no differences found at the sub-acute time point and so this data was not included in the current analyses (Coyle et al., 2022). Resting EEG and TMS-EEG data were processed and analysed offline using EEGLAB (Delorme et al., 2004), TESA (Rogasch et al., 2017) FieldTrip (Oostenveld et al., 2011) and custom scripts on the MATLAB platform (version R2017a). Please see Supplementary Materials for more detailed information on recording and preprocessing of the EEG and TMS-EEG data.

Resting EEG consisted of recording during eyes open (3 min) and eyes closed (3 min) conditions. For oscillatory power computation, EEG data were submitted to a frequency transformation based on fast Fourier transform using the ‘mtmfft’ method and Hanning taper (from 0.1 Hz to 100 Hz in steps of 0.2 Hz) to calculate the average power within four frequency bands: theta (4–8 Hz), alpha (8–12 Hz), beta (12–30 Hz) and gamma (30–45 Hz). Average total power was calculated across all epochs within each frequency band for each time point (sub-acute, T1, and T2) and condition (eyes open and eyes closed), resulting in a single value for each participant, within each frequency band and each time point and each condition at each electrode. In the sub-acute study (Coyle et al., 2022), cluster-based permutation analyses identified group differences for alpha power that were most pronounced in the right fronto-central region, generating a region of interest (ROI) for the current analyses. As only mean alpha power during eyes closed was found to be significantly different between groups at the sub-acute time point, this was the only resting EEG measure included in the current analyses. For statistical analysis of oscillatory power in the current study, non-parametric cluster based permutation statistics assessed differences between groups in eyes closed alpha at each time point separately. Electrodes in the alpha power ROI included; F2, FC2, FC4, FC6, CZ, C2, C4, C6, T8, CP2, CP4, CP6, P2, P8.

TMS-EEG involved administering 100 single TMS pulses with an inter-pulse interval of 4 seconds (with a 10% jitter). Intensity was 110% of resting motor threshold (RMT) and TMS was applied over the left dorsolateral prefrontal cortex (DLPFC) at the F3 electrode using the 10/20 system of placement. During TMS-EEG, participants listened to white noise through intra-auricular earphones (Etymotic Research, ER3-14A, USA) to limit the influence of auditory processing of the TMS click (Rogasch et al., 2014). The white noise sound level was adjusted for each participant until they reported that background noise was barely audible. TEP analysis focussed on four separate peaks known to occur following stimulation of the prefrontal cortex, the N45, P60, N100 and P200 (Rogasch et al., 2014; Rogasch et al., 2015). At the sub-acute time point mTBI participants demonstrated smaller (more negative) P60 and greater N100 TEP amplitudes (Coyle et al., 2022). Differences were most pronounced in the left parieto-occipital region for P60 and the right fronto-central region for N100, forming ROI’s for the current analyses. Following previous research, the N100 was defined as the amplitude averaged across the window between 0.09 and 0.135 s following the TMS pulse; while the P60 was identified as the mean amplitude occurring between 0.05 and 0.07s. For statistical analysis, non-parametric cluster-based permutation statistics assessed differences in mean amplitude between groups for each TEP component for each ROI. Electrodes in the P60 ROI cluster included; CP5, P7, P5, P3, P1, PO7, PO3, POZ, O1 and the N100 ROI cluster included; P7, P5, P3, P6, P8, PO7, PO3, POZ, PO4, PO8, O1, OZ, O2

### 2.3 Statistical Analyses

Statistical analyses were performed using R Studio (version 1.1.463) (Team RC., 2018). To investigate longitudinal change, whilst accounting for the effects of group and time, mixed linear models (MLM’s) were applied for each of our clinical, cognitive and neural outcome measures of interest. MLM’s are the recommended method of analysing longitudinal data in clinical samples (Garcia et al., 2017). They offer a flexible approach that allows for explicit modelling of the non-independence of repeated measures while accounting for missing data. The modelling was conducted including all participants who were tested in the sub-acute phase of the study using the *lme4* package (Bates et al., 2015) with the restricted maximum likelihood (REML) approach.

In the first instance, we fitted a maximal random effects structure (Barr et al., 2013), which included random intercept and slopes for participant by group and time. However, the full models did not converge, so a reduced model was used, modelling only the random intercept for participant. In the final model, all effects were taken as random at the participant level, and group and time estimates and statistics reported are at the population level. MLM assumptions of normality of residuals, linearity, homogeneity of variance were met. Although normality of outcome variable is not a MLM assumption, skewed distributions often result in a violation of the normality of residuals assumption, leading researchers to transform the outcome variable to improve the error distribution. *p* values for fixed effects regression coefficients were estimated via conditional F-tests using the *sJplot* package. Model significance (interaction and main effects) were evaluated using ANOVA (Type II Wald F tests) from the *car* package. Calculating *p* values for MLM’s is controversial, however the Kenward-Rogers approximation for degrees of freedom (df) is considered the best approach and was also utilised throughout (Luke et al., 2017). In addition to *p* values, fixed effect estimates and confidence intervals (CI) are included to aid interpretation. In instances where the outcome measure was required to be log transformed, geometric least square means were extracted and back transformed with *emmeans* (Lenth et al., 2018). Conditional R^2^, which includes variance explained by both fixed and random effects was included as a measure of variance explained.

Commonly, MLM are used to test hypotheses of effect modification in response to an intervention and post-hoc analyses are only conducted when an interaction effect is present. Interaction effects are of interest when trying to establish the effect of two (or more) factors on an outcome measure. However, the baseline value of the outcome variable can affect the validity of estimations of an interaction effect. For example, group differences at baseline that persist across time can result in a smaller quantity of change and present as a main effect of group and not a significant interaction effect. Due to these limitations, and our a priori aims, we also conducted exploratory post-hoc investigations of significant main effects as well as significant interaction effects.

In total 7 MLM’s were conducted. Clinical measures included mean total fatigue (MFI), mood (HADS) and post-concussion symptom severity (RPQ). The cognitive measure included was mean total words recalled for RAVLT list learning trial 1. Neural measures included; mean alpha power at right fronto-central ROI, P60 at parieto-occipital ROI and N100 amplitude at right fronto-central ROI. The models estimated the means, variances and co-variances of the random coefficients (participants) for the quantitative predictors (clinical, cognitive and neural outcome measures). The categorical predictors of group, time point and their interaction (group*time) were included as fixed effects. Due to violations of normality in specific outcome measures, natural log transformations were conducted for all clinical outcome measures and for alpha power. Log transformation can only be applied to positive data. For mean total mood and PCS the data included 0, resulting in a constant (log (y +1)) being added to all values. To assist interpretation when outcome variables were on log scale, coefficients and confidence intervals were back transformed and these values were used in tables and figures. This involves doing the opposite of the mathematical function used in the data transformation, in this case (exp(y)). Untransformed data was used for cognitive and TMS-EEG outcome measures.

## 3. Results

58 participants (30 mTBI) were enrolled between Feb 2017-March 2019. Of these, 48 (21 mTBI) completed 3-month follow-up and 40 (15 mTBI) completed 6-month follow-up assessments. See Supplementary Table 2 and Supplementary Figure 1 for more information on participants who completed each timepoint. Missing data per outcome measure and time point did not exceed 25% of the total. At baesline and 3-month follow-up, the groups did not differ significantly on measures of sex, age and pre-morbid intelligence (all *p* > .05), however controls had a higher level of education *(p* = .035). At 6-month follow-up, the control group were younger (*p* = 0.04) and had a higher level of education (*p* = 0.025).

### 3.1 Clinical and Cognitive Measures

For total mood symptoms, fixed effects of group and the interaction between group*3-month follow up significantly predicted total mood symptoms (*p* < 0.05). When evaluating model significance, a main effect of group [*F* _(1, 52)_ = 8.31, *p* = 0.006] and a trend group*time interaction effect [*F* _(2, 67)_ = 2.96, *p* = 0.058] were demonstrated. Post-hoc comparisons revealed that mTBI had significantly greater mood scores compared to controls (*p* <.001) at the sub-acute time point, but not at 3-month or 6-month follow up (See Figure 1A and Supplementary Table 3). The model explained 75.2% of the overall variance.

**Figure 1.**
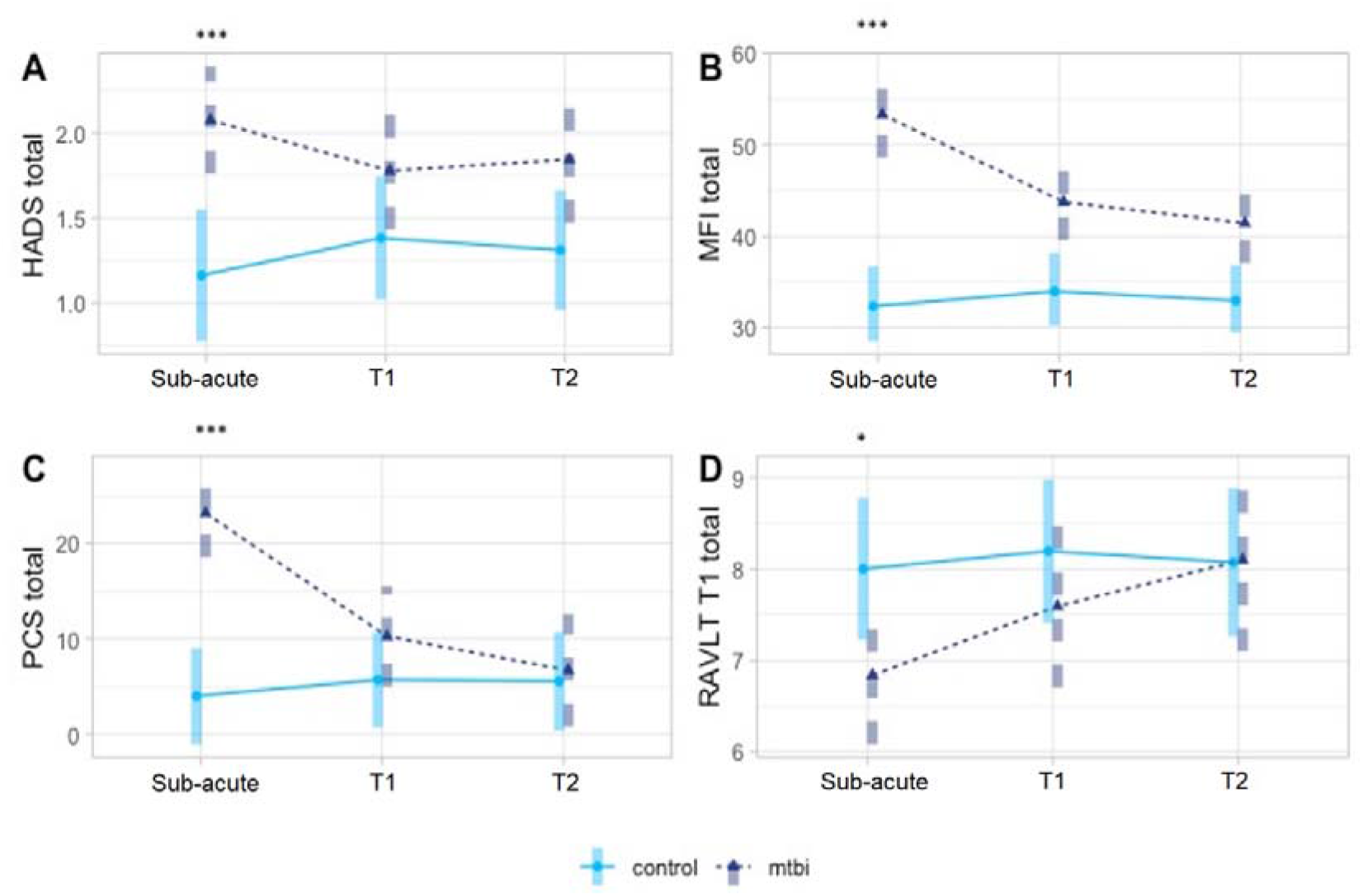
Model means by group (control = light blue, mTBI = dark blue) and time point for clinical measures and cognitive measures. A— Hospital Anxiety and Depression Scale (HADS). B — Multidimensional fatigue inventory (MFI). C — Post-concussive symptom (PCS) severity. D — RAVLT trial 1 (RAVLT T1). *** p < 0.001; ** p < 0.01; * p < 0.05. Bars indicate 95% confidence intervals.

In the total fatigue analysis, the fixed effects of group, the interaction between group*3-month and group*6-month were shown to significantly predict total fatigue (*p* < 0.05). When evaluating model significance, a main effect of group [*F* _(1, 43)_ = 28.17, *p* < 0.001], time point [*F* _(2, 60)_ = 5.56, *p* =.006] and interaction effect for group*time point [*F* _(2, 60)_ = 6.19, *p* =.004] were demonstrated. Post-hoc comparisons revealed that mTBI had significantly greater total fatigue scores at all time points compared to controls (*p* <.05) (See Figure 1B and Supplementary Table 3). The model explained 71.4% of the overall variance.

For post-concussion symptoms (PCS), the fixed effects of group and interaction between group*3-month and group*6-month follow up were shown to significantly predict total PCS (*p* < 0.05). When evaluating model significance, a main effect of group [*F* _(1, 53)_ = 6.65, *p* = 0.042], time [*F* _(2, 87)_ = 6.65, *p* = 0.002], and group*time interaction effect [*F* _(2, 89)_ = 14.65, *p* < 0.001] were demonstrated. Post-hoc comparisons revealed that mTBI had significantly greater PCS scores compared to controls at the sub-acute time point, but not at 3-month or 6-month follow up (See Figure 1C and Supplementary Table 3). The model explained 47.5% of the overall variance.

When investigating verbal learning performance (i.e. RAVLT T1), only the fixed effect of group was shown to significantly predict RAVLT T1 score (*p*< 0.05). When evaluating model significance, no main effect of group, time or interaction of group* time were demonstrated. For confirmation of sub-acute findings, exploratory contrasts were conducted, which confirmed that mTBI participants recalled fewer words than controls (*t* (97) = 2.11, *p* = 0.04) at the sub-acute time point (See Figure 1D and Supplementary Table 3). The model explained 45.6% of the overall variance.

MLM coefficients and 95% confidence intervals for fixed effects for clinical and cognitive outcome measures are included in Table 1. Model means (geometric means for log transformed outcome measures and estimated marginal means for untransformed outcome measures) are graphically displayed in Figure 1. Means, standard error (SE) and post-hoc comparisons are included in Supplementary Table 3.

**Table 1.**
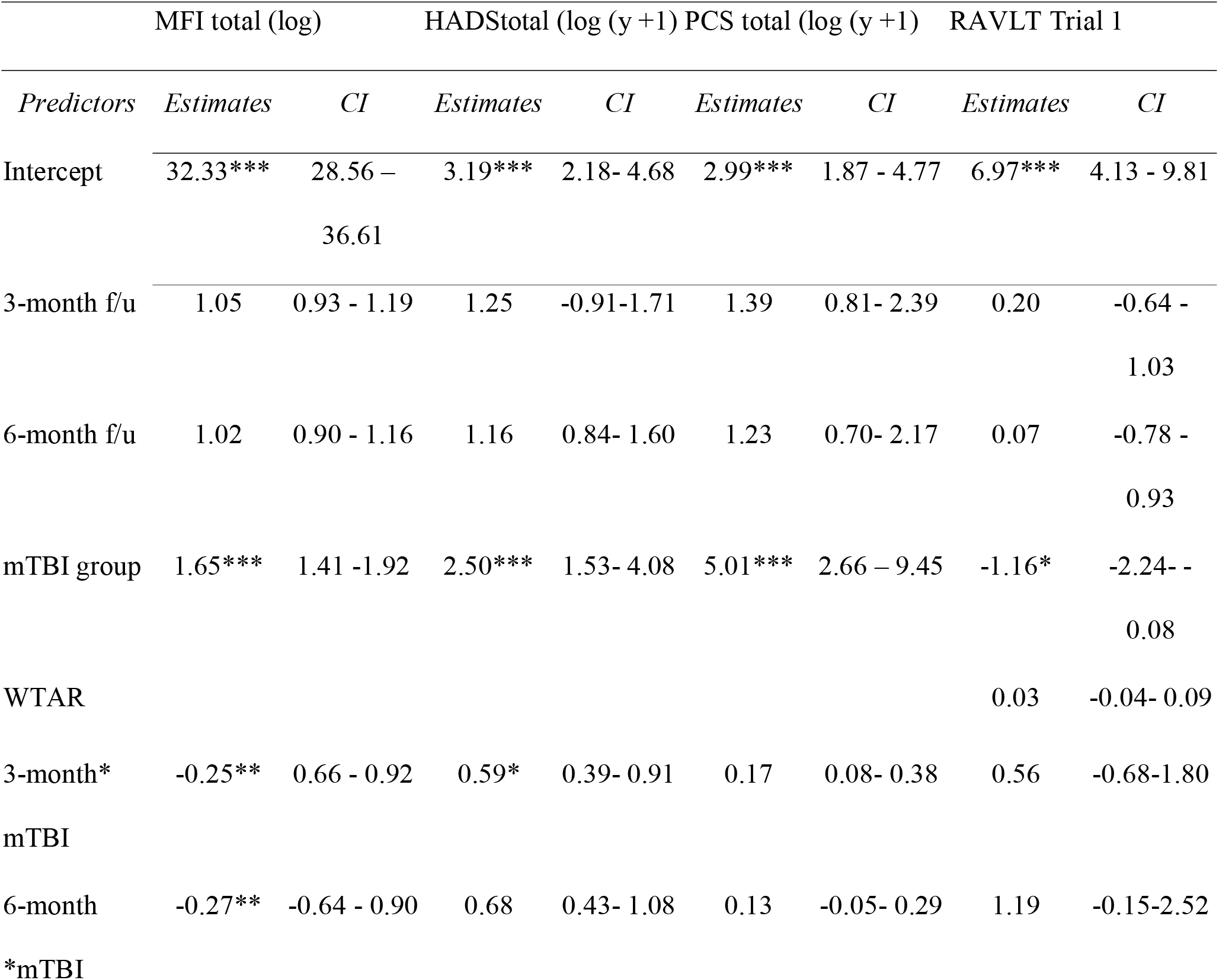
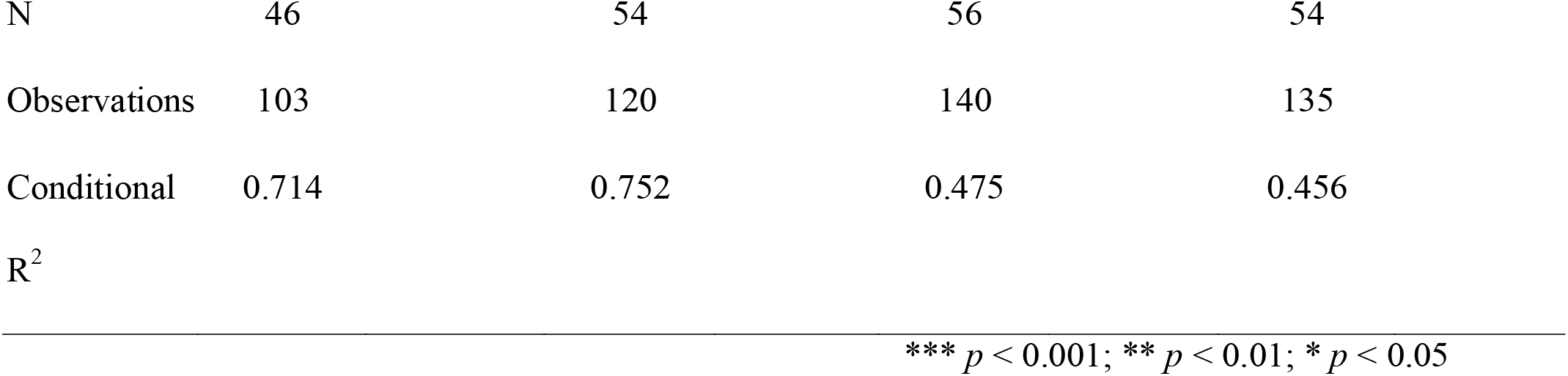
Regression coefficients and confidence intervals (CI) for each fixed effect from LMM’s for clinical and cognitive measures

|  | MFI total (log) |  | HADStotal (log (y +1) |  | PCS total (log (y +1) |  | RAVLT Trial 1 |  |
| --- | --- | --- | --- | --- | --- | --- | --- | --- |
| *Predictors* | *Estimates* | *CI* | *Estimates* | *CI* | *Estimates* | *CI* | *Estimates* | *CI* |
| Intercept | 32.33*** | 28.56 – 36.61 | 3.19*** | 2.18- 4.68 | 2.99*** | 1.87 - 4.77 | 6.97*** | 4.13 - 9.81 |
| 3-month f/u | 1.05 | 0.93 - 1.19 | 1.25 | -0.91-1.71 | 1.39 | 0.81- 2.39 | 0.20 | -0.64 - 1.03 |
| 6-month f/u | 1.02 | 0.90 - 1.16 | 1.16 | 0.84- 1.60 | 1.23 | 0.70- 2.17 | 0.07 | -0.78 - 0.93 |
| mTBI group | 1.65*** | 1.41 -1.92 | 2.50*** | 1.53- 4.08 | 5.01*** | 2.66 – 9.45 | -1.16* | -2.24- - 0.08 |
| WTAR |  |  |  |  |  |  | 0.03 | -0.04- 0.09 |
| 3-month*<br>mTBI | -0.25** | 0.66 - 0.92 | 0.59* | 0.39- 0.91 | 0.17 | 0.08- 0.38 | 0.56 | -0.68-1.80 |
| 6-month<br>*mTBI | -0.27** | -0.64 - 0.90 | 0.68 | 0.43- 1.08 | 0.13 | -0.05- 0.29 | 1.19 | -0.15-2.52 |

|  |  |  |  |  |
| --- | --- | --- | --- | --- |
| N | 46 | 54 | 56 | 54 |
| Observations | 103 | 120 | 140 | 135 |
| Conditional | 0.714 | 0.752 | 0.475 | 0.456 |
| R<sup>2</sup> |  |  |  |  |

\*\*\* $p < 0.001$ ; \*\* $p < 0.01$ ; \* $p < 0.05$

### 3.2 Neural Measures

#### 3.2.1 Resting EEG

For alpha power, the fixed effect of group and 3-month follow up were significant predictors of alpha power (p < 0.05) and the interaction between mTBI group*3-month follow up was at trend level (t (81) = -1.948, p = 0.055). When evaluating model significance, a main effect of time was demonstrated [F _(2, 80)_ = 14.11, p < 0.001]. Exploratory within group contrasts demonstrated reduced alpha power at 3 and 6 month follow up compared to the sub-acute time point for mTBI participants and reduced alpha power at 3 month follow up compared to the sub-acute time point for control participants (Supplementary Table 5). For confirmation of the sub-acute findings, exploratory between group contrasts demonstrated alpha power at the sub-acute time point was significantly greater for mTBI participants (See Figure 2A and Supplementary Table 4) The model explained 88% of the overall variance.

**Figure 2.**
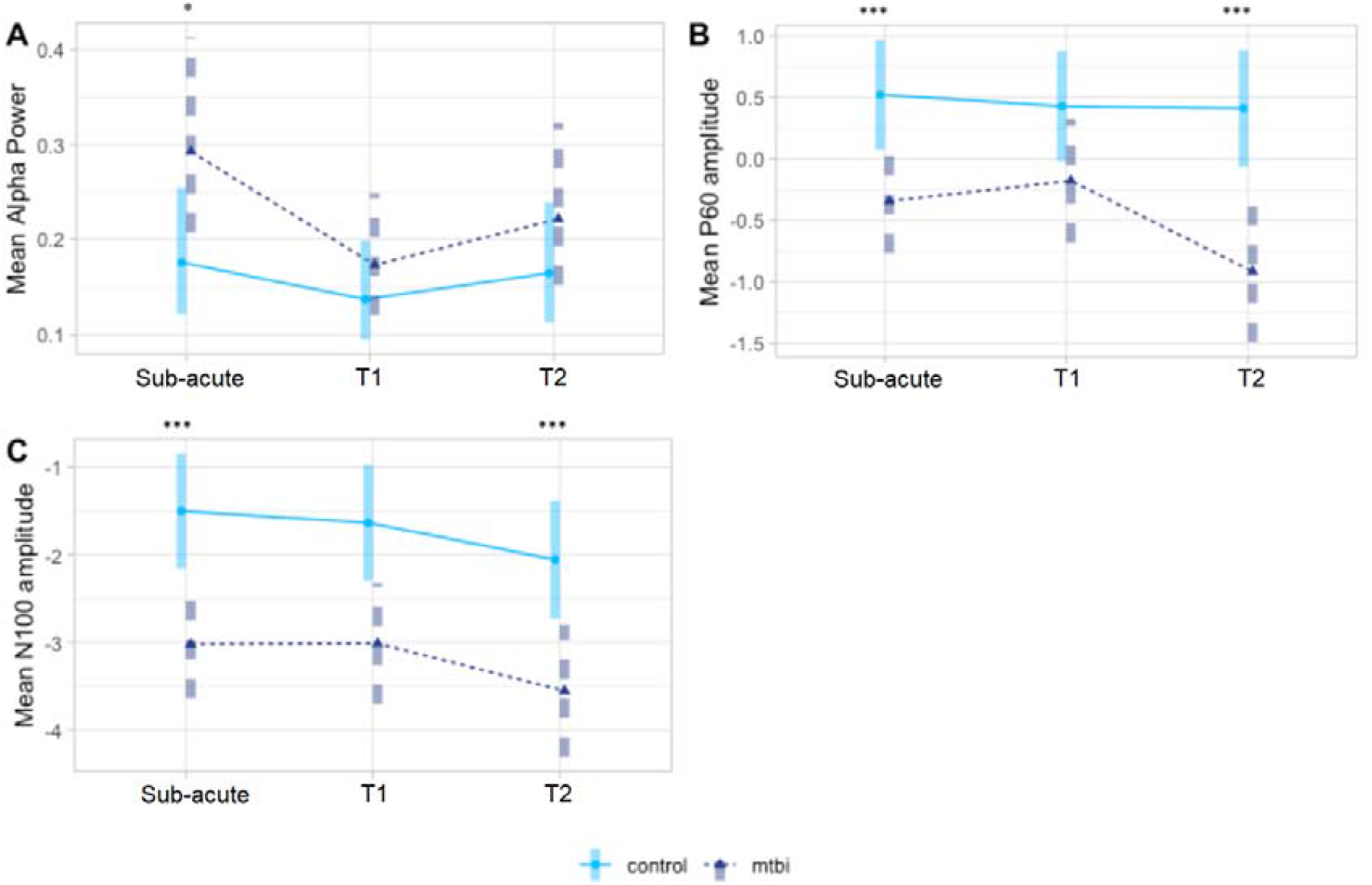
Model means by group (control = light blue, mTBI = dark blue) and time point for EEG and TMS-EEG measures. A—Average total alpha power in right fronto-central ROI B — Average P60 amplitude in left parieto-occipital ROI C— Average N100 amplitude in right fronto-central ROI. *** p < 0.001; ** p < 0.01; * p < 0.05. Bars indicate 95% confidence intervals.

#### 3.2.2 TMS-EEG

Single-pulse TMS over left dorsolateral prefrontal cortex (DLPFC) resulted in the expected characteristic series of negative and positive peaks. For the current analyses only P60 and N100 TEP components were investigated (determined by the differences detected at the sub-acute time point).

For P60 amplitude, the fixed effect of group was a significant predictor of P60 amplitude. Evaluation of model significance demonstrated a main effect of group (*F* _(1, 52)_ = 14.04, *p* < 0.001). Exploratory post hoc comparisons demonstrated that mTBI participants had smaller (more negative) P60 amplitude at the sub-acute time point and T2 compared to controls (See Figure 2C, Supplementary Table 4, and Supplementary Figure 2). The model explained 40% of the overall variance.

When investigating N100 amplitude, the fixed effect of group was a significant predictor of N100 amplitude. Evaluation of model significance demonstrated a main effect of time (*F* _(2, 82)_ = 3.29, *p* = 0.04) and group (*F* _(1, 53)_ = 13.24, *p* < 0.001) but no interaction of group*time (*p* > 0.05). Exploratory post hoc comparisons demonstrated that mTBI participants had greater (more negative) N100 amplitude at all time points (See Figure 2D and Supplementary Table 4). No within group differences were demonstrated (*p* > 0.05). The model explained 70.4% of the overall variance.

MLM coefficients and 95% confidence intervals for fixed effects for neural outcome measures are included in Table 2. Model means (geometric means for log transformed outcome measures and estimated marginal means for untransformed outcome measures) are graphically displayed in Figure 2. Means, SE and post-hoc comparisons are included in Table Supplementary Table 4.

**Table 2.**
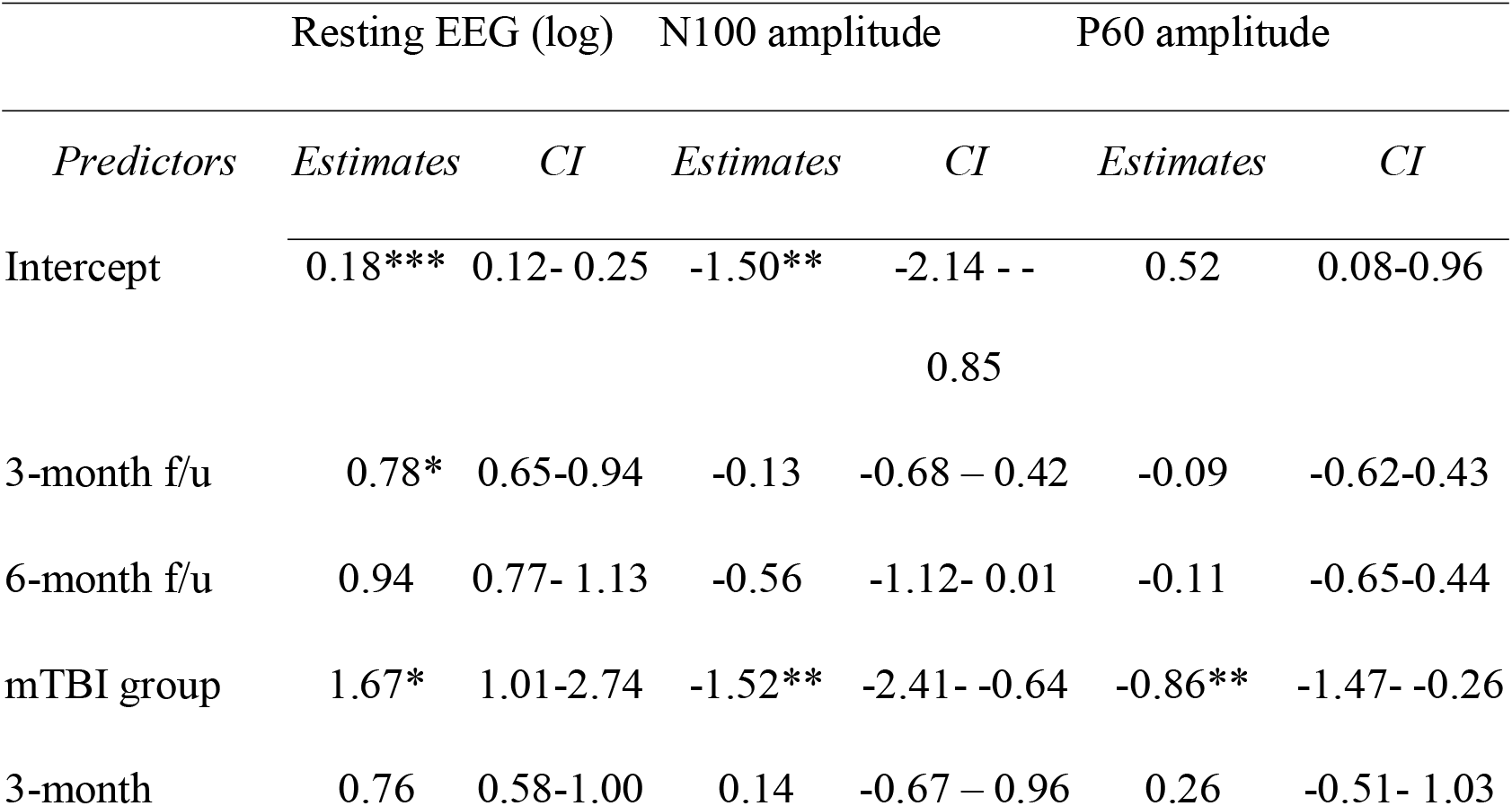
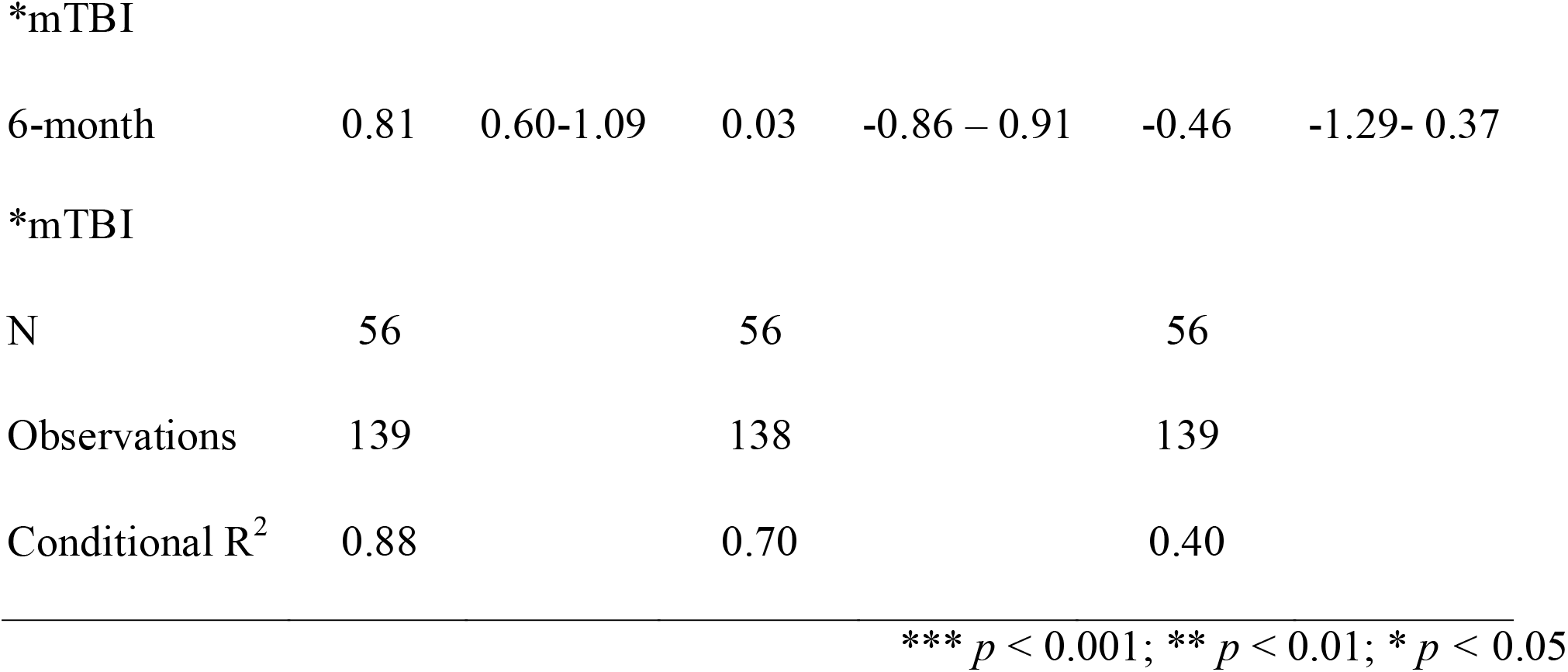
Regression coefficients and confidence intervals (CI) for each fixed effect from linear mixed effects regression models for neural measures

|  | Resting EEG (log) |  | N100 amplitude |  | P60 amplitude |  |
| --- | --- | --- | --- | --- | --- | --- |
| *Predictors* | *Estimates* | *CI* | *Estimates* | *CI* | *Estimates* | *CI* |
| Intercept | 0.18*** | 0.12- 0.25 | -1.50** | -2.14 - -<br>0.85 | 0.52 | 0.08-0.96 |
| 3-month f/u | 0.78* | 0.65-0.94 | -0.13 | -0.68 – 0.42 | -0.09 | -0.62-0.43 |
| 6-month f/u | 0.94 | 0.77- 1.13 | -0.56 | -1.12- 0.01 | -0.11 | -0.65-0.44 |
| mTBI group | 1.67* | 1.01-2.74 | -1.52** | -2.41- -0.64 | -0.86** | -1.47- -0.26 |
| 3-month | 0.76 | 0.58-1.00 | 0.14 | -0.67 – 0.96 | 0.26 | -0.51- 1.03 |
| $\ast_{\text{mTBI}}$ |  |  |  |  |  |  |
| 6-month | 0.81 | 0.60-1.09 | 0.03 | -0.86 – 0.91 | -0.46 | -1.29- 0.37 |

${}^*\text{mTBI}$
|  |  |  |  |
| --- | --- | --- | --- |
| N | 56 | 56 | 56 |
| Observations | 139 | 138 | 139 |
| Conditional $R^2$ | 0.88 | 0.70 | 0.40 |

\*\*\* $p < 0.001$ ; \*\* $p < 0.01$ ; \* $p < 0.05$

## 4. Discussion

The association between clinical and cognitive symptoms and neurophysiological changes following mTBI remains poorly understood. The current study used multimodal assessments in a longitudinal design to characterise recovery trajectories. The results showed that on measures of mood, post-concussion symptoms and verbal learning, impairments resolved within 3 months’ post-injury. However, greater symptoms of fatigue were reported by the mTBI group across all three time points. For the neural measures, the higher values of resting alpha power seen in the mTBI group at 4 weeks’ post-injury were resolved at 3 months with no further changes at 6 months. The P60 TMS-EEG measure at parieto-occipital electrodes demonstrated a similar pattern. Group differences, with mTBI participants demonstrating smaller (more negative) amplitude, were present at 4 weeks’ post injury, ameliorated at 3 months, and re-emerged at 6 months. The N100, demonstrated group differences across all three time points, with mTBI participants demonstrating greater (more negative) amplitude.

### 4.1 Clinical and Cognitive Symptom Recovery

Mood and PCS group differences were shown to resolve by 3 months’ post-injury, likely reflecting an amelioration of some acute neurophysiological changes (McKee et al., 2015). This is consistent with mTBI literature that reports most individuals experience partial or complete symptom resolution within a few weeks to 3 months’ post-injury (Rohling et al., 2011). However, our mTBI participants were not completely symptom free at 3 months, with greater fatigue reported and persisting to 6 months’ post-injury. Fatigue is a common complaint following neurological insult and has increasingly been recognised following mTBI (Chandhuri et al., 2004). A large longitudinal study reported 68% prevalence of fatigue at 1 week post mTBI, 38% at 3 months and 34% at 6 months (Norrie et al., 2010) and a cross-sectional study reported 33% of their sample experienced severe fatigue at 6 months’ post-injury (Stulemeijer et al., 2006; Stulemeijer et al., 2010). A possible explanation for persistent fatigue in our mTBI participants is presented by the pattern of results across the cognitive and neural measures. This will be discussed below. On a measure of cognitive function that involves information processing, the mTBI group showed impairments at 4 weeks. However, group differences were no longer present at 3 or 6 months’ post-injury, suggesting initial verbal learning acquisition deficits had recovered. This is consistent with a systematic review of meta-analyses demonstrating the majority of studies report a return to cognitive baseline by 90 days post-injury (Rohling et al., 2011).

### 4.2 Neural Measures Recovery

Following mTBI, neural measures obtained both while participants were at rest and during response to perturbation with TMS were different between the mTBI and control groups. However, recovery trajectories were somewhat variable across these different measures. mTBI participants’ alpha power differences at 4 weeks’ post-injury resolved at 3 and 6 month follow up time points. Within group comparisons revealed both groups demonstrated significant alpha power reductions from the first (sub-acute time point) to second session (3 month follow up). However, only mTBI participants demonstrated reductions between the first and third session (6 month follow up), supporting the between group findings that suggest greater alpha power normalises across recovery in mTBI. Alpha power is state and task dependent suggesting it may be vulnerable to environmental stimuli (Bazanova et al, 2012). Consequently, participants may have become accustomed to the novelty of the laboratory testing environment by the second testing session, resulting in reduced alpha power compared to the first session. Further research establishing the test–retest reliability of alpha power measures in mTBI and healthy controls would be of benefit.

TMS-EEG findings indicated mTBI participants demonstrated smaller (more negative) parieto-occipital P60 TEP amplitude at 4 weeks’ post-injury and at 6-month follow up. N100 amplitude was persistently larger (more negative) at all time points. To our knowledge this is the first study to assess TMS-EEG measures in the DLPFC in a longitudinal design in mTBI. P60 is considered a putative marker of excitation and although its functional significance is still being established, research has demonstrated that in response to a paradigm designed to induce inhibition, P60 and N100 modulations are positively correlated (Noda et al., 2017). The authors interpreted these findings as suggestive of an antagonistic or compensatory E: I relationship between the components that represents stable neurophysiological characteristics (Noda et al., 2017). However, other research has suggested that TEPs are significantly influenced by somatosensory and auditory aspects of TMS pulses, and as such the functional relevance of the P60 and N100 is still debated (Conde et al., 2019; Nikouline et al. 1999, Biabani et al. 2019, Biabani et al. 2021). Inspection of topoplots demonstrates the pattern of activation is comparable to that in previous research reporting P60 component topography following administration of TMS to the DLPFC (Chung et al., 2018a; Chung et al., 2018b). However, cluster based analyses in our sub-acute study demonstrated P60 group differences were largest in the right parieto-occipital region, which has not previously been reported (Coyle et al., 2022). Furthermore, mTBI participants demonstrated a negative potential during the P60 latency window (see Supplementary Figure 2). This may reflect dysregulated excitation; however, the paucity of previous research suggests our P60 findings should be interpreted with caution. Alternatively, our N100 findings are consistent with longitudinal studies by Miller et al., (2014) and Edwards et al., (2017) which reported changes related to inhibitory function in MEPs. As the N100 has been suggested to at least partly reflect measure of GABA_B_ mediated inhibition and dysregulation is suggestive of changes to GABA-ergic synaptic transmission (Premoli et al. 2014; Rogasch et al. 2013, Rogasch et al. 2015, Belardinelli et al. 2021, Kaarre et al. 2018, Noda 2020).

Magnetic resonance spectroscopy (MRS) has also suggested the N100 reflects a balance between GABA-inhibitory and glutamate-excitatory levels (Du et al., 2018), however using multi-modal tools to investigate the neurobiological mechanisms associated with TEP components warrants further investigation. Cross-sectional research examining TMS-EEG over the DLPFC on average 5 years post-injury also supports our finding of persistent alterations, with greater N100 amplitude being demonstrated in mTBI participants (Tallus et al., 2013). It has previously been proposed that changes to inhibition may be long-term compensatory mechanisms against glutamate excitotoxicity following mTBI (De Beaumont et al., 2012). However, smaller P60 and greater N100 in mTBI suggests the balance is dysregulated in favour of greater inhibition, with potential implications for efficient information processing. Alternatively, it is also possible that differences in the P60 and N100 reflect somatosensory or auditory processing differences (Conde et al., 2019; Nikouline et al. 1999, Biabani et al. 2019, Biabani et al. 2021). In summary, the current TEP findings demonstrate altered neural reactivity to TMS pulses following mTBI that persists at 6 months’ post-injury. This suggests neurophysiological recovery may be more prolonged than is able to be captured by standard neuropsychological measures.

The TMS-EEG findings suggest persistent altered neural responses, significant differences from healthy controls still being demonstrated 6 months’ post mTBI. These changes may reflect alterations to the finely balanced excitation and inhibition of the brain, which is essential for cortical circuit function, and imbalance is thought to lead to inefficient or impaired cognitive function (He et al., 2019; Legon et al., 2015). These findings coincide with the persistently elevated fatigue reported by the participants in our mTBI group, in the presence of normalised cognitive function, and future research should seek to explore these associations further.

The resolution of changes in resting brain activity by 3 months is also of interest. Previous research has hypothesised that compensatory mechanisms during recovery are indicative of the adaptive capacity of neural systems (Tallus et al., 2013; Maruishi et al., 2007; Dean et al., 2015). The brain’s capacity to recover and adapt highlights the importance of investigating post-injury plasticity to develop novel approaches to therapeutic intervention. Neuroplasticity mechanisms also offer a possible explanation for variability in our findings across time points, mTBI pathophysiology and compensatory activity uniquely interacting across recovery.

### 4.5 Limitations

Several study limitations should be noted. Firstly, there were challenges related to sample size and characteristics. Attrition bias through the systematic loss of participants across time is a significant complicating factor in most longitudinal clinical research. Our choice of statistical analysis, mixed linear models, mediated information loss by including all available participant data and as such this analysis was more robust to missing data. The possibility that individuals who dropped out may have differed systematically from those who completed all sessions was explored. Analyses demonstrated individuals who did not return for 3 month follow up did not differ on injury severity (as measured by GCS), total mood, fatigue or PCS at the sub-acute time point from those who completed all sessions. Attrition also contributed to a reduced sample size at the 6-month follow up (*N*=15 in the mTBI group), preventing us from conducting subgroup analyses, such as investigating asymptomatic vs symptomatic mTBI participants. Regarding sample characteristics, the control group had a higher level of education than the mTBI group. Although we included pre-morbid IQ as a fixed effect for MLM verbal learning (RAVLT) performance analyses, this difference may have mediated cognitive performance. Furthermore, although alternative forms were used at each time point for cognitive tasks assessing memory, there is the possibility that practice effects may have influenced results for other cognitive measures. Secondly, the study design does not allow us to establish causal mechanisms. Although repeated measures provide information on change over time post-injury, it is possible that group differences may have preceded injury. Thirdly, the application of TMS could be optimised. We used the F3 scalp location as an approximate landmark for the DLPFC, over which single pulse TMS was administered. Although this method has been shown to provide a relatively accurate estimate of the DLPFC (Rusjan et al., 2010) future research utilising MRI-based neuro-navigational software could help to further increase accuracy when targeting this region. Accurate source localisation analyses using individual MRI templates could also improve our understanding of the neuroanatomical bases of our findings.

### 4.6 Future Directions

The current findings provide a foundation for future research utilising multimodal investigations to explore recovery in mTBI. Although we have demonstrated the utility of TMS-EEG measures as potential biomarkers for injury and recovery, further research concurrently acquiring information from additional modalities is needed. For example, haemodynamic, microstructural and biochemical changes are relevant following mTBI, highlighting the potential application of concurrent functional magnetic resonance imaging (fMRI), diffusion-weighted or diffusion-tensor imaging (DWI, DTI) and magnetic resonance spectroscopy (MRS) across recovery. A multimodal approach will assist with establishing the functional significance of TEP components, clarifying their diagnostic and prognostic utility and linking changes in excitation and inhibition to functional and structural connectivity. It is also necessary to conduct investigations with larger sample sizes to comprehensively explore variability in recovery trajectories. Lastly, future research should aim to extend the current study by identifying specific treatment targets and therapeutic windows. Neuromodulation for clinical and therapeutic applications to promote recovery in mTBI has previously been proposed and the present findings offer a significant step forward in this regard (Hoy et al. 2019; Li et al., 2015). Longitudinal multimodal approaches with sufficient power will improve our understanding of optimal neural targets and therapeutic windows and potentially help to identify individuals at risk of persistent symptoms.

## 4.7 Conclusions

These findings suggest mTBI dysregulates the dynamic equilibrium that underpins efficient neuronal communication and greater effort is required to maintain function. Substantially contributing to our understanding of neurophysiological changes across recovery post mTBI, our study highlights that on sensitive neurophysiological and fatigue measures, persistent alterations are present at 6 months’ post-injury. Variability across recovery may also be reflective of the interaction between mTBI pathophysiology and compensatory activity. Comprehensively characterising the multifaceted nature of recovery across time has significant potential to optimise the development of interventions. Future research should aim to improve our understanding of temporally optimum windows and therapeutic targets and link functionally relevant neuroplastic changes to persistent symptoms.

## Data Availability

We do not have ethical approval to make this data publicly available, as our approval predated our inclusion of such approvals (which we now do routinely).

## Acknowledgements

We would like to thank Caley Sullivan (Research Officer, Epworth Centre for Innovation in Mental Health, Monash University) for their assistance with scripts for data analysis and Eldho Paul (Research Fellow, Department of Public Health and Preventative Medicine, Monash University) for their assistance with mixed linear model development. This work was supported by an Australian Postgraduate Award Scholarship (HLC) and a National Health and Medical Research Council Fellowship (1135558) (KEH).

