## Supplemental Materials for "Recovery of clinical, cognitive and cortical activity measures following mild traumatic brain injury (mTBI): a longitudinal investigation"

### **Supplementary Information**

### **Materials and Methods**

###### *Participants*

Supplementary Table 1. Clinical characteristics of mTBI group

| Variables | *N* = 30 |
| --- | --- |
| Days post injury (M/SD/range)) | 19.70/ 16.96/ 10-31 |
| Short (0-10 days post) | 6.7% |
| Medium (11-20 days post) | 46.6% |
| Long (21-31 days post) | 46.6% |
| Amnesia |  |
| Retrograde | 33.3% |
| None | 30% |
| Retrograde and anterograde | 23.3% |
| Unknown | 13% |
| GCS |  |
| 15 | 53.3% |
| 14 | 20% |
| 13 | 3% |
| unknown | 23.3% |
| LOC (%) |  |
| Yes | 40% |
| No | 43.3% |
| Unknown | 16.7% |
| PCS severity |  |
| Mild | 30% |
| Moderate | 36.7% |
| Severe | 10% |
| None | 23.3% |
| History of mTBI (%) |  |
| Yes | 56.7% |
| No | 43.3% |
| CT pathology (%) |  |
| Yes | 43.3% |
| haematoma | 23.3% |
| haemorrhage | 6.7% |
| haematoma & haemorrhage | 6.7% |
| haematoma & fracture | 6.7% |
| No | 53.3% |
| Unknown | 3.3% |
| Mechanism of Injury (%) |  |
| Fall from bicycle | 43.3% |
| Fall | 20% |
| Head strike by object | 13.3% |
| Pedestrian vs. motor vehicle | 13.3% |
| Motor vehicle accident | 9.9% |
| Other injury at time of accident |  |
| Yes | 63.3% |
| No | 36.7% |

Supplementary Table 2. Demographic Information for Control and mTBI across time points

|  | Control | mTBI |  | p | d |
| --- | --- | --- | --- | --- | --- |
| Baseline |  |  |  |  |  |
| N | 26 | 30 |  |  |  |
| sex = male (%) | 18 (69.2) | 23 (76.7) | *χ2* = .11 | 0.746 | 0.09 |
| age (mean (sd)) | 31.65 (9.06) | 35.43 (10.31) | *t* = -1.46 | 0.154 | 0.39 |
| education (mean (sd)) | 16.94 (2.59) | 15.25 (3.19) | *t* = 2.19 | 0.035* | 0.58 |
| WTAR (mean (sd)) | 41.76 (5.63) | 38.00 (7.86) | *t* = 1.91 | 0.053 | 0.51 |
| 3 month follow up |  |  |  |  |  |
| N | 26 | 21 |  |  |  |
| sex = male (%) | 18 (69.2) | 17 (81%) | *χ2* = .33 | 0.562 | 0.17 |
| age (mean (sd)) | 31.65 (9.06) | 36.05 (11.25) | *t* = -1.45 | 0.155 | 0.43 |
| education (mean (sd)) | 16.94 (2.59) | 14.55 (2.42) | *t* = 3.27 | 0.002* | 0.95 |
| WTAR (mean (sd)) | 41.76 (5.63) | 37.58 (7.63) | *t =* 1.91 | 0.064 | 0.59 |
| 6 month follow up |  |  |  |  |  |
| N | 24 | 15 |  |  |  |
| sex = male (%) | 17 (70.8%) | 13 (86.7%) | *χ2* = .56 | 0.453 | 0.24 |
| age (mean (sd)) | 31.08(9.11) | 38.87(11.15) | *t* = -2.27 | 0.032* | 0.76 |
| education (mean (sd)) | 16.9 (2.69) | 14.7(2.6) | *t* = 2.53 | 0.016* | 0.83 |
| WTAR (mean (sd)) | 41.83 (5.12) | 39 (5.49) | t = 1.53 | 0.139 | 0.53 |

Supplementary Table 3. Numbers of participants and completed assessments

|  | Control (n) | mTBI (n) |
| --- | --- | --- |
| Time point |  |  |
| Sub-acute | 28 | 30 |
| 3-month f/u | 27 | 21 |
| 6-month f/u | 25 | 16 |

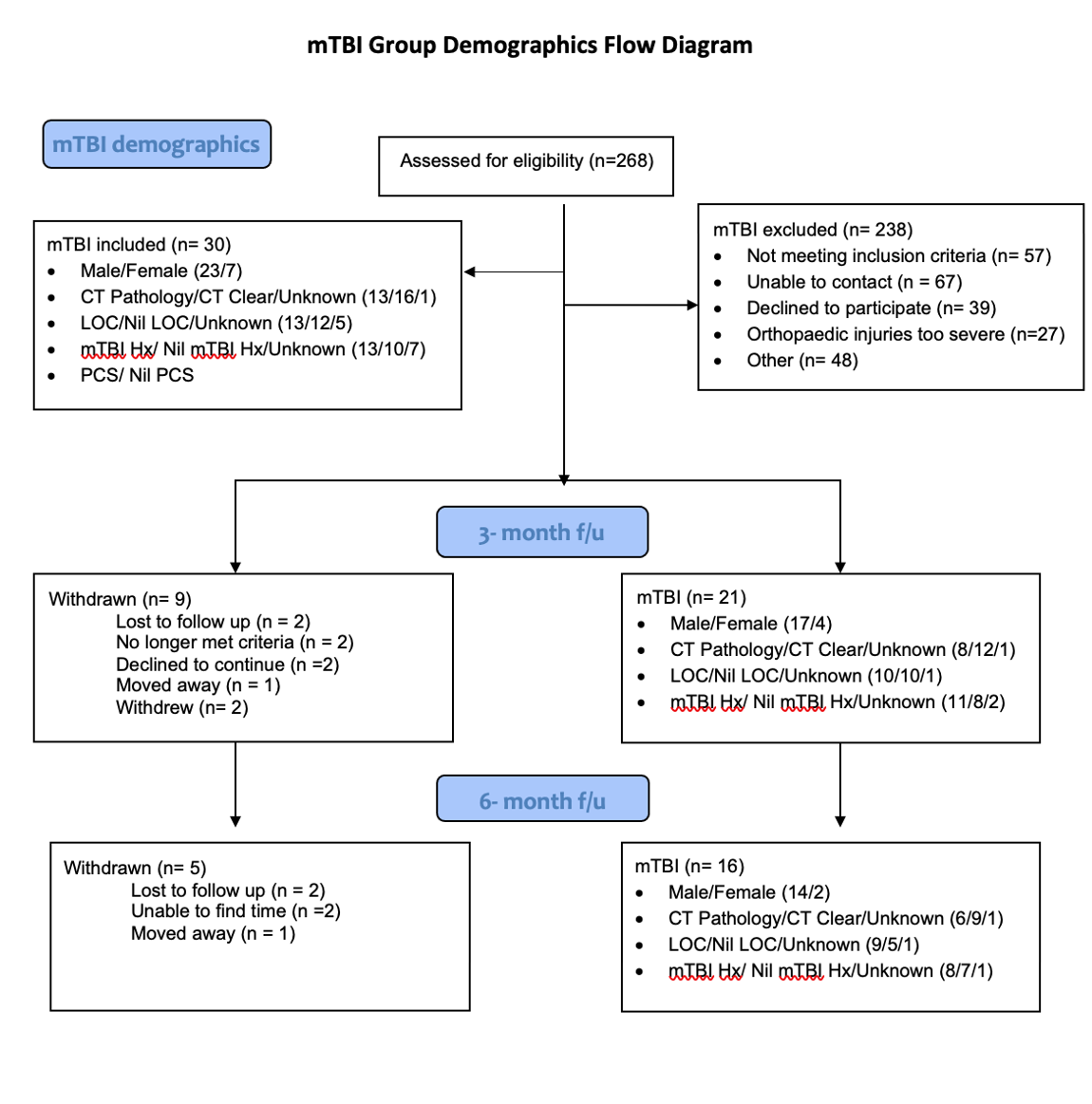

Supplementary Figure 1. CONSORT flow diagram displaying mTBI demographics and reasons for attrition across time points.

*Measures*

###### EEG Recording

EEG was recorded with TMS-compatible Ag/AgCl electrodes and a DC coupled amplifier (SynAmps2, EDIT Compumedics Neuroscan, Texas, USA). Fifty electrodes were used from a 64-channel Easycap EEG cap (AF3, AF4, F7, F5, F3, F1, Fz, F2, F4, F6, F8, FC5, FC3, FC1, FCz, FC2, FC4, FC6, T7, C5, C3, C1, Cz, C2, C4, C6, T8, CP5, CP3, CP1, CP2, CP4, CP6, P7, P5, P3, P1, Pz, P2, P4, P6, P8, PO7, PO3, POz, PO8, PO4, O1, Oz, O2). Electrodes were referenced on-line to CPz and grounded to FPz. All data was recorded with a high acquisition rate (10,000Hz) and low-pass filtered (DC- 2,000 Hz) using a large operating window (200 mV). Electrode impedances were kept below 5 kΩ throughout the recording. Participant instructions prior to resting EEG, and task related stimuli were presented during EEG using Presentation® software (Version 18.0, Neurobehavioral Systems, Inc., Berkeley, CA, [www.neurobs.com)](about:blank).

###### Resting EEG Pre-processing

Resting EEG data were down sampled (500 Hz), bandpass filtered (fourth-order, zero-phase, Butterworth filter, 0.1-100 Hz), bandstop filtered (48-52 Hz; to remove 50Hz line noise) and epoched into two second epochs. Automatic artefact rejection was completed, which first checked if more than 3% of epochs included electrodes that varied by more than -250 to 250 microvolts and excluded those electrodes. Next epochs were excluded if they showed a variation of more than 5 SD’s of kurtosis for individual channels, or 3 SD’s for all channels. Lastly, epochs with power within the frequencies 25 to 45Hz that exceeded -100 or 30 dB were excluded (power in these frequencies usually reflects muscle activity). Manual artefact rejection was then completed to ensure the automatic process did not miss significant artifacts, the data being visually inspected to remove epochs with excessive noise (i.e. muscle artefact), and bad channels (i.e. disconnected). Independent component analysis (Fast-ICA algorithm using the ‘tanh’ contrast function) decomposition were then applied to the data.

Rejected electrodes were re-constructed using spherical interpolation (Perrin et al. 1989), and data was re-referenced to the average reference. Resting EEG data was then split into two files (at the marker for the auditory tone), creating separate files for eyes open (EO) and eyes closed (EC) conditions.

###### TMS-EEG Pre-Processing

TMS-EEG data were epoched around the TMS pulse (-1000ms to 1000ms) and baseline corrected to the pre-TMS pulse period (-500ms to -50ms). Data around the large signal from the TMS pulse (-5 to 15ms) were removed and linearly interpolated. Data were downsampled from 10,000Hz to 1,000Hz. An initial round of independent component analysis (Hyvarinen and Oja 2000) (FastICA) was then performed to remove components containing any large residual TMS-evoked EMG artefacts. A bandpass filter (1–100 Hz) was then applied and line noise was removed using a bandstop filter (48–52 Hz). Data was again visually inspected and any remaining noisy epochs removed. Finally, a second round of FastICA was performed to eliminate any remaining components representing blink, decay and noise-related artefacts. Both rounds of component rejection following FastICA utilised a semi-automated artefact detection algorithm, based on a previous research (Rogasch et al. 2014) and using TESA toolbox as a guide (Rogasch et al. 2017). Components representing the following artefacts were removed; eye blinks and saccades (mean absolute z score of the two electrodes larger than 2.5), persistent muscle activity (high frequency power that is 60% of the total power), decay artefacts and other noise-related artefacts (one or more electrode has an absolute z score of at least 4).

#### **Results**

Supplementary Table 4. Geometric means and standard error (SE) for clinical measures, estimated means and standard error for cognitive measures.

| *MFI total* | Control | mTBI | *p* |
| --- | --- | --- | --- |
| Sub-acute | 32.33 (2.05) | 53.33 (2.49) | <0.001 |
| 3-month | 33.96 (1.99) | 43.74 (2.20) | 0.002 |
| 6-month | 32.95 (1.83) | 41.44 (2.32) | 0.005 |
| *HADS total* |  |  |  |
| Sub-acute | 4.14 (1.37) | 9.35 (1.17) | 0.005 |
| 3-month | 4.92 (1.35) | 7.75 (1.23) | 0.125 |
| 6-month | 4.33 (1.32) | 7.49 (1.28) | 0.090 |
| *PCS total* |  |  |  |
| Sub-acute | 4.00 (2.54) | 23.23 (2.33) | <0.001 |
| 3-month | 5.71 (2.51) | 10.28 (2.66) | 0.21 |
| 6-month | 5.56 (2.61) | 6.71 (2.98) | 0.77 |
| *RAVLT T1* |  |  |  |
| Sub-acute | 7.99 (0.39) | 6.84 (0.38) | 0.04 |
| 3-month | 8.20 (0.40) | 7.59 (0.44) | 0.32 |
| 6-month | 8.07(0.41) | 8.09 (0.50) | 0.97 |

Supplementary Table 5. Geometric means and standard error (SE) for neural measures

| *Alpha Power* | Control | mTBI | *p* |
| --- | --- | --- | --- |
| Sub-acute | 0.18 (0.04) | 0.29 (0.05) | 0.04 |
| 3-month | 0.13 (0.03) | 0.17 (0.03) | 0.37 |
| 6-month | 0.16 (0.03) | 0.22 (0.04) | 0.27 |
| *P60 amplitude* |  |  |  |
| Sub-acute | 0.52 (0.23) | -0.34 (0.21) | <0.01 |
| 3-month | 0.43 (0.22) | -0.17 (0.25) | 0.07 |
| 6-month | 0.41(0.03) | -0.91 (0.28) | <0.01 |
| *N100 amplitude* |  |  |  |
| Sub-acute | -1.49 (0.32) | -3.02 (0.30) | <0.01 |
| 3-month | -1.63 (0.33) | -3.01 (0.34) | <0.01 |
| 6-month | -2.05 (0.34) | -3.55 (0.38) | <0.01 |

Supplementary Table 6. Within group contrasts for alpha power

| *Control* | t. ratio | p |
| --- | --- | --- |
| Sub-acute/T1 | 2.63 | 0.27 |
| Sub-acute/T2 | 0.67 | 0.78 |
| T1/T2 | -1.85 | 0.16 |
| *mTBI* |  |  |
| Sub-acute/T1 | 5.0 | <0.01 |
| Sub-acute/T2 | 2.37 | 0.05 |
| T1/T2 | -2.04 | 0.11 |

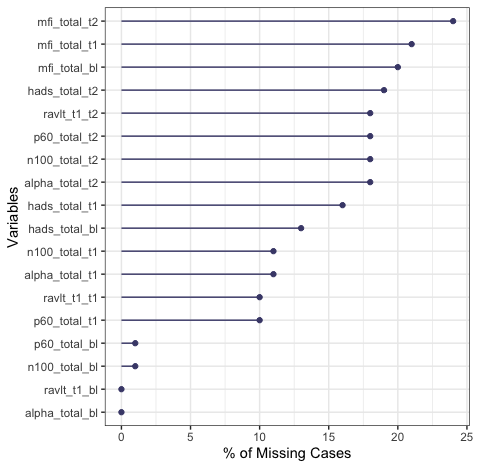

Supplementary Figure S1. Percentage of missing data per outcome measure and time point.

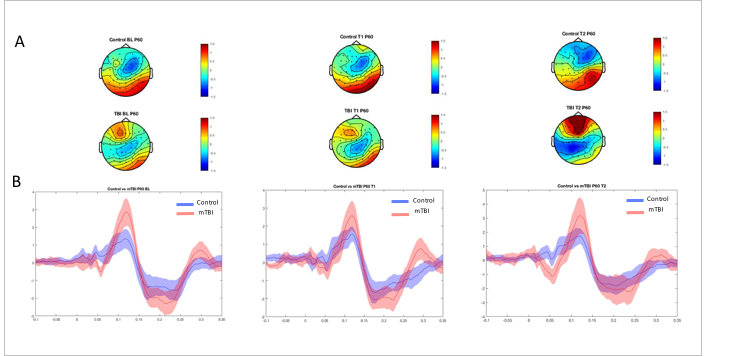

**Supplementary Figure 2**. A. Topoplots demonstrating the distribution of activity in each group within the P60 component latency range from 50- 70 ms post TMS pulse. B. TEP waveforms from electrodes in the left parieto-occipital region (the ROI identified from cluster based analyses in our baseline study, Coyle et al 2022).
